# The CAPP 2 Study Protocol: Strengthening the capacity of healthcare providers to reduce the impact of COVID-19 on African, Caribbean, and Black communities in Ontario

**DOI:** 10.1101/2023.08.11.23293990

**Authors:** Josephine Etowa, Hugues Loemba, Liana Bailey, Sanni Yaya, Charles Dabone, Egbe B. Etowa, Bishwajit Ghose, Wale Ajiboye, Jane Tyerman, Marian Luctkar-Flude, Jennifer Rayner, Onyenyechukwu Nnorom, Robin Taylor, Sheryl Beauchamp, Goldameir Oneka, Bagnini Kohoun, Wangari Tharao, Haoua Inoua, Ruby Edet, Joseph Kiirya, Soraya Allibhai, Ky’okusinga Kirunga, Janet Kemei

## Abstract

**Introduction:** The COVID-19 pandemic emerged as an unprecedented challenge for healthcare systems across the world disproportionately impacting immigrant and racialized populations. Canadian African, Caribbean, and Black (ACB) communities representing some of the most vulnerable populations in terms of their susceptibility to health risks, receipt of adequate care, and chances of recovery. The COVID-19 ACB Providers Project (CAPP 2) aims to strengthen the ability of health care providers (HCP) to address this community’s COVID-19 related healthcare needs. Informed by CAPP 1.0 Project, a mixed-method study which examined COVID-19 pandemic impact on ACB communities in Ontario (Ottawa and Toronto), this second study seeks to develop and implement educational programs on five key areas (modules) to strengthen the capacity of HCPs working with ACB populations. The five modules (topics) include: 1) COVID-19 and its impacts on health, 2) social determinants of health and health inequities, 3) critical health literacy, 4) critical racial literacy, and 5) cultural competence and safety.

**Methods and analysis:** An implementation science approach will guide the development, implementation, and evaluation of the evidence-informed interventions. Intersectionality lens, socio-ecological model (SEM) and community-based participatory research (CBPR) frameworks will inform the research process. To ensure active stakeholder engagement, there will be a Project Advisory Committee comprised of 16 ACB community members, health providers, and partner agency representatives. Five modules will be developed: two virtual simulation games in collaboration with leading simulation experts, and three non-simulation modules.

**Ethics and dissemination:** Ethics approval was granted by the University of Ottawa Research Ethics Board on July 18th, 2023 (H - 01-23 - 8069). The results of this study will be disseminated in community workshops, an online learning platform, at academic conferences and in peer-reviewed publications.

## Strengths and limitations of this study

- Provides innovative methods for the development of health equity programs for diverse providers.
- Proposes methods of evaluating different educational methods to determine efficiency of programs.
- Measures of programme ‘success’ will be based on participant experience rather than clinical measures.
- Adopts tenants of community based participatory research to integrate significant program change.

## Background

The COVID-19 pandemic emerged as an unprecedented challenge for healthcare systems across the world. Globally, the risk and burden of COVID-19 has been disproportionately higher among immigrant and racialized populations [1–5]. Canadian African, Caribbean and Black (ACB) communities represent some of the most vulnerable populations in terms of their susceptibility to health risks, receipt of adequate care and chances of recovery [6]. Black communities in Canada experience higher rates of COVID-19 related infections and deaths [7]. Existing data shows that the neighbourhoods with the highest proportions (≥ 25%) of ‘racialized’ groups, or visible minorities, experience a COVID-19 mortality rate twice as high as neighborhoods with the lowest proportions (< 1 %) of racialized populations [8].

Social determinants, including structural inequalities and discrimination, are known to account for the disproportionate health risks and differential health outcomes experienced in ACB populations [9–11]. In the case of COVID-19, excess cases and deaths have been attributed to disproportionately high rates of co-morbid conditions (e.g., diabetes, hypertension) in these communities [12]. Contributing structural factors include income, employment, food insecurity, and the built environment [13–14]. These conditions may for example manifest themselves as risky working conditions outside the home and needing to use public transportation. In Canada, data is limited but we know from national surveys that Black Canadians exceed 50% on economic vulnerability arising from the COVID-19 crisis. During the pandemic, 61% saw a decrease in their income, 50% had difficulty meeting their financial obligations and 47% were unable to pay their mortgage or rent on time [15]. The increased burden of COVID-19 morbidity and mortality among ACB populations has translated into greater challenges for health systems and for governments through loss of social capital, productive labour force, and erosion of cultural equity [16–17].

Evidence suggests that ‘one size does not fit all’ in terms of the response to public health disasters in vulnerable populations [18–19]. It is well-documented that ACB communities experience multiple and intersecting barriers to accessing appropriate and responsive health services. These barriers include institutional discrimination, poor representation in healthcare leadership, in research and in decision-making, and lack of awareness of available services as well as lack of culturally responsive services in relevant languages, and competent health professionals [20–21]. Numerous scholars have identified the urgent need to build capacity, as well as reduce stigma and paternalism among health providers working with ACB communities [22–23]. In the absence of these factors, inequalities are magnified, and scapegoating persists, with discrimination remaining long after [24]. There is also widespread acknowledgement that ACB communities and scholars need to be involved in all aspects of prevention, treatment and outreach [25–26]. As such, this project proposes an innovative way of utilizing high quality and real-time evidence to address contextual vulnerability and challenges experienced by ACB communities.

This project will address five specific themes stemming from data from the CAPP 1 project [27] with the aim of educating healthcare providers. In 2020-21, this project’s predecessor, the COVID-19 ACB Providers Project (CAPP) 1 aimed to examine the challenges experienced by ACB communities and identify strategies to build providers’ capacity to address their COVID-19 related health outcomes. This mixed methods research project generated four priority areas to ensure that health and service providers meet the needs of ACB communities to: 1. Transform Health System’s Capacity for Anti-Black Services, 2. Strengthen ACB Community’s Capacity for Authentic Participation in Health Policy, Practice Research, Education, and Leadership, 3. Create Programs and Policies that Foster Disaggregated Race-based Data Collection, Governance, Develop Strategic Partnerships, and 4. Create Accountability Measures for Anti-Black Racist Healthcare within all Levels of Healthcare System. This COVID-19 ACB Providers Project (CAPP) 2 aims at meeting these priority areas by creating capacity-building interventions focused on addressing the key themes highlighted in the CAPP predecessor project. More specifically this CAPP 2 will create interventions (modules) focused on the following five key themes or topics: 1) COVID-19 and its impacts on health, 2) social determinants of health and health inequities, 3) critical health literacy, 4) critical racial literacy, and 5) cultural competence and safety.

To create engaging and interactive learning resources, particularly with the onset of the COVID-19 pandemic, the education sector has adopted and utilised webinars more extensively [28]. The term “webinar” refers to communication over the internet between two or more participants using audio, video and interactive activities such as polls and quizzes, with possibilities to allocate participants into breakout rooms to work together on a topic or solve a problem [29]. Webinars have been used both synchronously and asynchronously as engaging learning formats. In synchronous format, learners participate together at a pre-defined time while in asynchronous format, they may engage webinar independently at any time. Evidence for the effectiveness of online training using webinars is growing [30–31]. In a recent systematic review, Gegenfurtner & Ebner [32] found that synchronous webinars were slightly more effective than control conditions (online asynchronous learning management systems and offline face-to-face classroom instruction), but these differences were trivial in size. Moreover, little research to date has examined the implementation and impact of online strategies to address professional capacity-building needs to reduce the impact of COVID-19 on ACB communities.

### Main Goals and Objectives

The main objective of this project is to accelerate the use of high quality and real-time evidence collected on the contextual vulnerability and challenges experienced by ACB communities to develop, implement and evaluate community-driven solutions to structural inequalities including systemic racism that continue to hamper the response and recovery from COVID-19. The specific objectives are to:

1. Engage ACB communities and health stakeholder groups in the development of community-driven strategies to reduce COVID-19 related health inequities.
2. Develop and implement innovative evidence-based health educational interventions (webinars) to reduce COVID-19 risk and burden in ACB communities during and after the pandemic.
3. Determine the reach and effectiveness of online capacity-building interventions (webinars) offered in various formats for healthcare providers.
4. Improve evidence-based communication among health stakeholder groups (i.e., health planners, health providers and ACB communities) to reduce COVID-19 related health inequities.

To determine the reach and effectiveness of online capacity-building interventions offered in various formats for healthcaThe re providers as indicated in objective 3 above, the following research questions will be answered:

1. What factors influence health care providers’ uptake of tailored synchronous and asynchronous capacity building interventions (i.e., critical racial literacy modules)?
2. How can tailored critical health and racial literacy educational program enhance health care service providers’ ability to provide effective healthcare for ACB people?

## Methods and Analysis

### Conceptual Frameworks

Community-based research frameworks and implementation science will be used to guide the development, implementation, and evaluation of the evidence-informed interventions to transform health system’s capacity to address anti-Black racism in healthcare. This approach involves ACB communities in the co-design and implementation of interventions, practice and policy tools. Community members will participate and take ownership of the process and co-create innovative solutions, in partnership with other stakeholders and decision makers. In addition, intersectionality lens, and socio-ecological model (SEM) frameworks will inform the research process. Intersectionality is an analytical tool used in equity work to understand and interpret the complexity of the world around us [33]. It emphasizes that health issues are influenced by broader social factors and do not occur in isolation, but rather these factors intersect and mutually-enhance their negative impacts on health outcomes [34]. SEM identifies a vast array of layered macro-, meso- and micro-level factors to consider when addressing health inequities including COVID-19 infection and its impact among racialized populations [35]. The Consolidated Framework for Implementation Research (CFIR) will be used to examine assess domains associated with effective implementation (i.e., intervention characteristics, outer setting, inner setting, characteristics of individuals, and process) [36]. CFIR is intended to be flexible in its application so that evaluators can tailor the framework to the specific program design, factors, and context being applied or studied. The focus of this project will be on constructs of the outer setting, such as external policy and incentives, and constructs of the inner setting, such as climate and leadership. The CFIR framework is a good fit for CAPP 2 specifically given its extensive use in community-based programs and interventions as well as its capacity to tailor interventions according to the research objectives. A variety of tools are being developed for the evaluation including website analytics, pre/post surveys, and follow-up interviews with providers and agencies.

### The Research Process

This 3-year research project will be integrative and organized in phases. Phase 1 will focus on pre-planning and module development - stakeholder engagement and knowledge mobilization tools development (year 1) to raise awareness among ACB people about the social determinants of COVID-19 vulnerabilities and to set the stage for the learning modules implementation and uptake; phase 2 innovative module development and implementation (year 2) for service providers collective empowerment and capacity building to build requisite skills, abilities and critical perspectives for effective COVID-19 and health equity responses among ACB communities; and phase 3, module implementation and evaluation as well as other knowledge mobilization activities (year 3) to mobilize providers, ACB people and other stakeholders in the collaborative development and implementation of evidence-informed programming, research and policy to address COVID-19-related health inequities in ACB communities.

**Phase 1:** To ensure active stakeholder engagement, a Project Advisory Committee (PAC) has been formed comprised of 16 ACB community members, health providers, partner agency representatives and other stakeholder groups. The PAC will meet quarterly for the duration of the project. The development of the online learning intervention will be led by J.E. and L.B. and assisted by the PAC and High-Impact Field-based Interventions (HIFI) laboratory. The intervention will consist of a series of five virtual educational modules for health providers aimed at: 1. Social determinants of health and health inequities; 2. COVID-19 and its impacts on health, 3. Critical health literacy; 4. Critical racial literacy; 5. Cultural competency and safety.

These modules will be grounded in current behavioral insights including the five communication principles; format messages in clear and easy to understand language, use social media platforms commonly used by ACB communities, use compelling stories from the community, make educational events social, and use evidenced-informed messages delivered by trusted messengers such as ACB community health professionals. An e-learning expert and instructional design consultants have been contracted to guide the development of the non-simulation modules for themes 1 and 2. The other three modules for themes 3, 4 and 5 will be virtual educational modules. Virtual simulation educational modules are innovative and accessible resources that provide effective instruction and critical thinking in healthcare education comparable with or exceeding outcomes associated with other simulation modalities or teaching methods [37–39]. The benefits of virtual simulation (VS) pedagogy were especially highlighted during the COVID-19 pandemic when most healthcare education had to shift to virtual formats. Learners report that VS is an engaging and enjoyable educational strategy that is easy to use and contributes to their learning [40]. The advantages of VS learning include the ability to place all learners in decision-making roles, to increase accessibility to larger groups of learners, to provide instant and evidence-based feedback, to increase flexibility in pacing, and have the potential to create a more psychologically safe learning environment [41]. Authors M.L-F. and J.T. are virtual simulation experts who will support the creation of these simulation modules using the Canadian Alliance of Nurse Educators using Simulation (CAN-Sim) virtual simulation design process [42]. The contracted e-learning specialist will lead the development of an accessible website to host the five learning modules.

**Phase 2:** Two different synchronous and asynchronous formats will be employed and tested for the implementation of capacity-building activities among health providers. The first format will target health providers in five pre-specified practice settings (i.e., Ottawa Public Health, Somerset West CHC, Southeast Ottawa CHC, AIDS Committee of Ottawa, African Caribbean Council on HIV in Ontario (ACCHO). In each setting the research team will work with local health partners to engage and deliver training to at least 75% of agency staff. Health Providers more broadly will also be targeted to engage in the asynchronous online modules housed in an interactive website developed for this project. The website and online modules will be promoted to both health care facilities (e.g., primary care, hospitals, LTC, community organizations) and provider groups (e.g., physicians, nurses, allied health staff). Dissemination will also be promoted through existing collaborations with organizations such as CAN-Sim. A minimum of 100 participants will be reached. Figure 1 illustrates the logic model [40] which will be used to track project indicators throughout.

**Figure 1.**
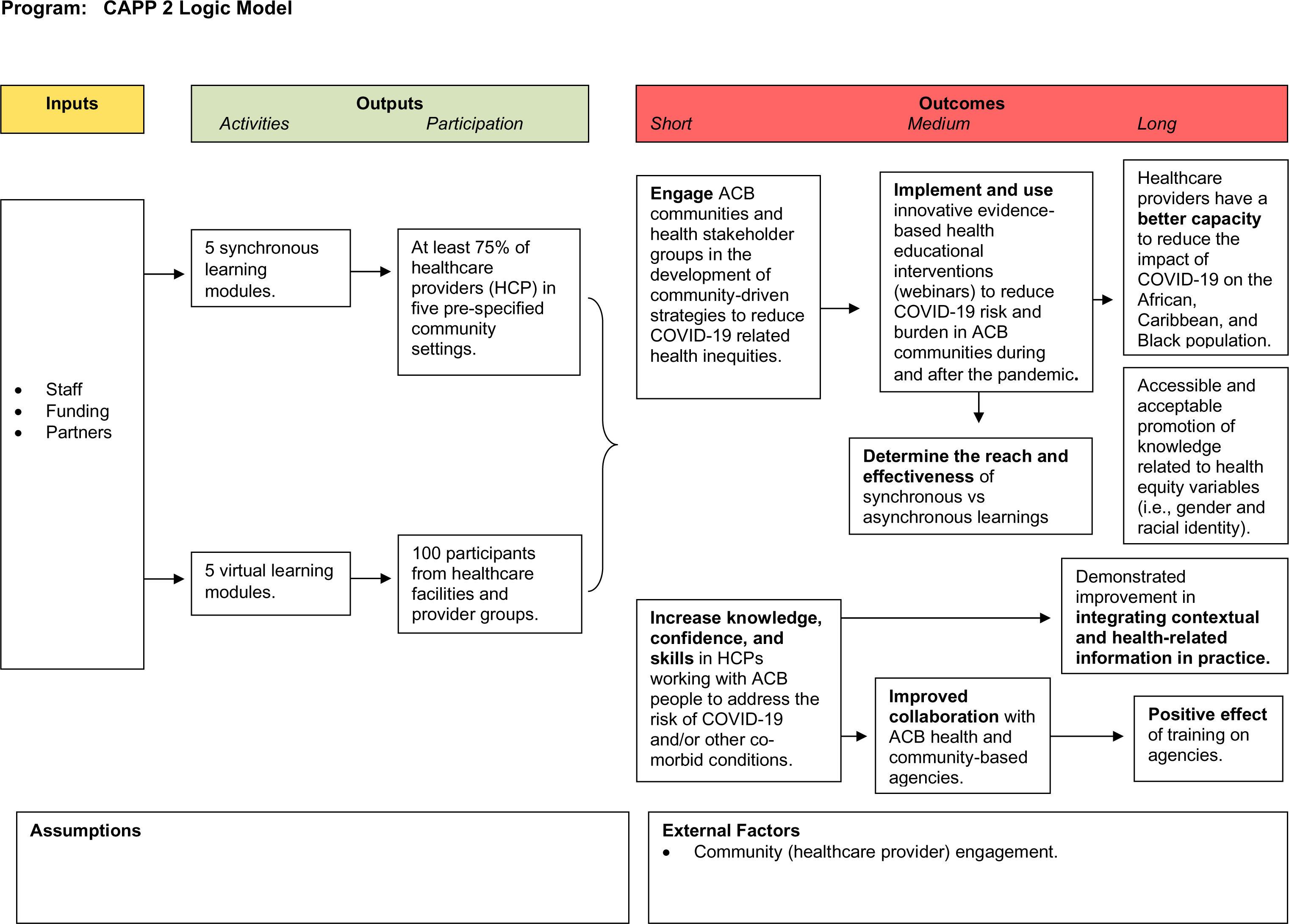
CAPP 2 Logic Model.

**Phase 3:** A comprehensive evaluation will be continuously conducted to assess the project’s activities and impacts, including implementation outputs, short-term and long-term outcomes (as seen in the above Logic Model). The implementation research will focus on the five domains found in the CFIR: intervention characteristics, outer setting, inner setting, characteristics of the individuals involved, and the process of implementation [36]. Further, this project will focus on the characteristics of the inner and outer settings that influence implementation (e.g. structural characteristics, policies, implementation climate, etc.). Specifically, this project is interested in learning whether health providers who participate in setting-specific training using the online modules are more likely to benefit from the training and/or engage in quality improvement/system transformation initiatives in their agencies than health providers who complete the online modules independently. The expected short-term outcome for providers includes increased knowledge, confidence, and skills in working with ACB people to address the risk of COVID and/or other co-morbid conditions. Long-term outcomes include a demonstrated improvement in integrating contextual and health-related information in practice, positive effect of training on agencies, and improved collaboration with ACB health and community-based agencies. Focus group discussions and demographic questionnaires have been developed (see supplemental materials) and will be provided to consenting participants at this phase.

Data analysis will be guided by the intersectionality lens and SEM frameworks. All focus group discussions (FGDs) will be audio-recorded and transcribed verbatim. We will use NVivo software for data management, storage and to facilitate analysis. Braun and Clarke’s [43] six-step thematic analysis process will guide the data interpretation and analysis. A theme is defined as a pattern found in the information that describes and organizes the possible observations, or interpretations of phenomena identified in the data [43]. This process will begin with the development of a code list informed by questions from the FGD guides and a systematic approach that involves: (1) familiarizing with the data; (2) generating initial codes; (3) developing a coding tree to guide the coding of transcripts; (4) identifying themes; (5) reviewing, defining and naming themes; 6) interpreting the narratives and stories; and (7) producing the report – a concise, coherent, logical, and non-repetitive account supported by vivid examples [43]. These iterative processes associated with qualitative analysis will ensure that preliminary interpretations are challenged, and that data is revisited in the light of further data collection and new insights into the data. During data analysis, preconceptions and assumptions will be challenged, and consensus will be reached in understanding the data. Thematic analysis is suitable for this study because it is participatory and accessible; it enhances participation among team members (knowledge users, peer research associates, and academic researchers) in collaborative data analysis and interpretation. Themes derived from the qualitative analysis will help to inform the evaluation of the online learning modules for healthcare providers.

### Patient and Public Involvement

Consistent with the CBPR principles interwoven throughout this study, the protocol has been developed collaboratively with community partners, knowledge users, and (community and academic) stakeholders since its conception. Research questions and objectives were formulated based off of community experiences, specifically those made evident by this project’s predecessor study CAPP 1 [27]. CAPP 1 exposed the gaps in care that this project aims to help fill. Specifically, this project exposed the five module themes. Community partner agencies have been selected to support the use of the five modules to train and recruit their staff (as described in the logic model in Figure 1) for this program evaluation. Collaborating organizations, such as CAN-Sim, will also help support dissemination through to their community of learners. A formal local advisory group has been created and will meet quarterly following project commencement to ensure meaningful involvement of all partners in decision-making related to study activities.

### Rigor, Ethics and Dissemination

To establish the trustworthiness and scientific rigor of this research, the research process will be guided by f the framework for assessing the quality of qualitative research postulated by Lincoln and Guba [44]; that is, credibility, transferability, confirmability, and dependability. Credibility refers to the confidence readers have in the analytical and interpretive processes and findings. In this research, transparent analytical steps are identified and informed by established qualitative research principles. Transcripts will be carefully verified and checked. In the analysis process, attention will be paid to misrepresentation of the evaluation component, focus group discussion (FGD) data. We will interrogate the data until data saturation is reached. Furthermore, the research team is trained in the conduct of ethical and sensitive research with this study populations. Transferability is the extent to which the study results might be relevant to similar populations that have similar characteristics. Transferability is dependent on “thick description”, which in this work will be the detailed and precise narrative that will be constructed following the comprehensive analysis of the evaluation data. The narrative description will provide sufficient detail for others to make a judgement on the quality of the results. Confirmability will be achieved by reflexive team and project advisory committee (PAC) meetings. Dependability will be achieved by the establishment of a clear audit trail and accurate documentation of the research processes and procedures, including the analytical process, field notes, digital recordings and transcripts.

Ethics approval was obtained (July 2023) from the Research Ethics Boards of the University of Ottawa, and operational sites permission will be obtained where necessary prior to commencing research. Voluntary written informed consent will be obtained from all participants. As part of the consenting process, participants will be assured that they do not have to answer every demographic or focus group discussion (FGD) question, they can choose to be recorded on audio (or not), and they can withdraw at any time. They will especially be informed that their non-consent will not affect their employment conditions. The principles of confidentiality and anonymity will be observed at all times, including in the storage of research materials. If instances of distress are encountered, participants will be offered information about counselling support services. The post training survey and focus group discussion will be performed in a secured digital platform or in-person location at the University of Ottawa or partner agency.

Accessible resources and practice/policy tools will be developed to enable health policymakers and planners throughout the province to meet the healthcare needs of ACB communities currently and post-pandemic. The wide-spread impact of COVID-19 demands that previously considered local concerns now become global concerns. Provincial collaboration such as the one in this project will bring different research perspectives to bear on the issue of COVID-19 and health service provision from the perspective of ACB people in Ontario. Specifically, this project will promote and strengthen knowledge exchange and links to providers, knowledge users and researchers across Ontario with the potential for national scale-up. It will facilitate exposure to innovative ideas and approaches. It will provide opportunities for redesigning and identifying promising models for provider capacity building informed by critical and diverse perspectives. While there are no quick solutions, sustained gains in providers’ COVID-19-related health equity capacity, such as the one that would be provided by this project, will successfully harness healthcare providers who can address critical post-pandemic health priorities.

## Authors’ Contributions

All authors drafted the manuscript. J.E. conceived the study and all investigators assisted with refining the research design, while collaborators assisted with the strategic assembly of team composition. Graduate students and other trainees assisted with proposal review, organization, editing and referencing, and overall formatting. All these categories of team members are authors and they all critically reviewed and revised the manuscript. All authors read and approved the final manuscript. Individual contributions are listed below.

Josephine Etowa: conceptualized the idea and drafted protocol.

Hugues Loemba: discussed initial ideas, brainstormed to refine ideas and research design. Liana Bailey: discussed initial ideas, brainstormed to refine ideas and research design.

Contributed to the draft protocol.

Sanni Yaya: discussed initial ideas, brainstormed to refine ideas and research design. Charles Dabone: discussed initial ideas, brainstormed to refine ideas and research design. Egbe B. Etowa: discussed initial ideas, brainstormed to refine ideas and research design. Bishwajit Ghose: discussed initial ideas, brainstormed to refine ideas and research design. Wale Ajiboye: discussed initial ideas, brainstormed to refine ideas and research design.

Jane Tyerman: discussed initial ideas, brainstormed to refine ideas and research design.

Marian Luctkar-Flude: discussed initial ideas, brainstormed to refine ideas and research design. Jennifer Rayner: discussed initial ideas, brainstormed to refine ideas and research design.

Onyenyechukwu Nnorom: reviewed proposal and informed team composition, and community partners.

Robin Taylor: discussed initial ideas, brainstormed to refine ideas and research design. Sheryl Beauchamp: assisted with proposal review, organization, editing and references, and overall formatting.

Goldameir Oneka: assisted with proposal review, organization, editing and references, and overall formatting.

Bagnini Kohoun: reviewed proposal and informed team composition, and community partners.

Wangari Tharao: reviewed proposal and informed team composition, and community partners. Haoua Inoua: reviewed proposal and informed team composition, and community partners.

Ruby Edet: reviewed proposal and informed team composition, and community partners. Joseph Kiirya: reviewed proposal and informed team composition, and community partners. Soraya Allibhai: reviewed proposal and informed team composition, and community partners. Ky’okusinga Kirunga: reviewed proposal and informed team composition, and community partners.

Janet Kemei: reviewed proposal and informed team composition, and community partners.

## Supporting information

Focus Group Discussion Guide

Demographic Survey

## Data Availability

All data produced in the present study are available upon reasonable request to the authors.

## Acknowledgments

The authors wish to thank all key stakeholders, partners and social/health services providers who have given their support to this study. The authors would further like to acknowledge that this work was presented at the Canadian Association of Schools of Nursing Biennial Conference in May of 2023.

