## Supplementary material for "The CAPP 2 Study Protocol: Strengthening the capacity of healthcare providers to reduce the impact of COVID-19 on African, Caribbean, and Black communities in Ontario": Focus Group Discussion Guide

**APPENDIX 3**

**Focus Group Discussion (FDG) Guide**

### Strengthening the capacity of healthcare providers to reduce the impact of COVID-19 on African, Caribbean, and Black communities in Ontario-Phase 2 (CAPP 2.0)

**Name of Interviewer:**

**Date:**

Hello, my name is________ and I am a __________. I will be conducting a focus group discussion (FGD) to debrief on your experiences with the 5-module series you completed on strengthening the capacity of healthcare providers to reduce the impact of COVID-19 on African, Caribbean, and Black (ACB) communities in Ontario. If it is okay with you, I will be recording our meeting and the recording will be transcribed. The purpose of recording and transcribing is so that we can have an attentive conversation. I assure you that all your comments will be confidential, and a pseudonym will be used on all information.

1. Describe ways that you found (self-directed online modules/ webinar modules) effective in helping you understand new information.
   If applicable (prompts):
   a) What were the benefits/ disadvantages of "real-time" learning?
   b) What were the benefits/ disadvantages of self-directed learning?
   c) What were the benefits/ disadvantages of the virtual simulation?

2. Please describe how the format of the modules was beneficial in improving your knowledge on:
   a) Social determinants of health and health inequities.
   b) Critical health literacy.
   c) Critical racial literacy.
   d) Organizational and provider cultural competency and safety.

e) COVID-19 and its impacts on health.

3. Are there are additional comments you would like to add that could improve the learning modules and their format to improve your knowledge and skills regarding increasing vaccine uptake within the most affected groups, such as the ACB community?

That concludes our interview. Thank you for participating in our study, your input greatly appreciated.
