## Supplementary material for "The CAPP 2 Study Protocol: Strengthening the capacity of healthcare providers to reduce the impact of COVID-19 on African, Caribbean, and Black communities in Ontario": Demographic Survey

**APPENDIX 4**
**Service Providers Demographic Form**

**Strengthening the capacity of healthcare providers to reduce the impact of COVID-19 on African, Caribbean, and Black communities in Ontario-Phase 2 (CAPP 2.0)**

**Background information**

- District of practice ________________________________
- Name of health facility _________________________

| Position / expertise | Years in role | Age | Sex | Ethnicity | Highest level of Education achieved | Ethical consent: verbal (V) or written (W) |
|---|---|---|---|---|---|---|
